# Global and local genetic overlap among ME/CFS, irritable bowel syndrome and psychiatric traits: a hypothesis-generating analysis

**DOI:** 10.64898/2026.06.08.26355171

**Authors:** Jun Hyun Lee

## Abstract

**Background:** Myalgic encephalomyelitis/chronic fatigue syndrome (ME/CFS) and irritable bowel syndrome (IBS) frequently co-occur following infection, yet shared genetic architecture at the locus level has not been systematically characterised.

**Aims:** To estimate global and local genetic correlations between ME/CFS (including infection-onset subgroup), IBS, major depressive disorder (MDD) and loneliness/isolation, and characterise ME/CFS cell-type heritability enrichment.

**Method:** GWAS summary statistics: DecodeME (15,579 ME/CFS; 9,738 infection-onset), FinnGen R9 (9,296 IBS), PGC MDD Wave 2 (45,396) and UK Biobank loneliness (N=455,364). LDSC for global correlations; LAVA for local correlations across 2,495 loci; MAGMA for cell-type enrichment (Descartes Human atlas); coloc.abf for colocalisation.

**Results:** All pairwise global correlations were significant after Bonferroni correction, including ME/CFS-all–MDD (r_g_=0.598, 95% CI 0.46–0.74) and ME/CFS-all–IBS (r_g_=0.573, 0.39–0.75). Of 4,232 local tests, 16 reached FDR<0.05; two loneliness×MDD loci were Bonferroni-significant. ME/CFS–MDD showed three FDR-significant local correlations, but all were boundary-estimated and non-Bonferroni-significant. A borderline infection-onset ME/CFS–IBS signal occurred at chr12q24.22 (ρ=1.000, FDR=0.046), but colocalisation did not support a shared causal variant (PP.H4=0.007). ME/CFS heritability was enriched in inhibitory neurons (P=1.2×10^−7^) and enteric nervous system neurons (FDR=0.004), with no FDR-significant peripheral immune cell-type enrichment in the atlas used.

**Conclusions:** High global ME/CFS–MDD correlation was accompanied by limited, boundary-estimated, non-Bonferroni-robust local sharing; the data do not support reducing ME/CFS to depression at the genetic-architecture level. Neural enrichment, including enteric nervous system neurons, supports involvement of neural components in ME/CFS susceptibility without excluding immune mechanisms. A borderline ME/CFS–IBS signal at a *NOS1*-containing region generated hypotheses requiring replication.

## 1. Introduction

Myalgic encephalomyelitis/chronic fatigue syndrome (ME/CFS) is a debilitating condition characterised by post-exertional malaise, cognitive impairment, and autonomic dysregulation, with an estimated global prevalence of 0.2–0.4%.^1^ A substantial proportion of cases are triggered by infections, including SARS-CoV-2, Epstein–Barr virus and enteroviruses.^2,3^ Irritable bowel syndrome (IBS) shares this post-infectious pattern, with 10–25% of cases arising after acute gastrointestinal infection.^4^ Epidemiological studies report two-to fourfold higher IBS prevalence in people with ME/CFS than in the general population,^5^ suggesting possible shared pathobiology.

The first large-scale ME/CFS GWAS identified eight genome-wide significant loci with enrichment in brain tissue expression rather than immune tissue,^2^ while IBS GWAS analyses have characterised overlapping genetic architecture with psychiatric and pain traits.^6^ Formal identification of shared local genetic loci between these co-occurring conditions has not, however, been performed. Global genetic correlations (r_g_) quantify aggregate polygenic overlap but do not identify specific shared regions or distinguish causal from pleiotropic mechanisms. Local genetic correlation analysis using LAVA^7^ tests for correlated genetic effects within discrete genomic windows, providing locus-level resolution; combined with single-cell transcriptomic cell-type enrichment, this approach links genetic architecture to biological substrates. We conducted a systematic cross-trait analysis of ME/CFS (total and infection-onset), IBS, MDD and loneliness, with the aims of (i) estimating genome-wide genetic correlations, (ii) identifying local genetic correlations, (iii) characterising the pattern of global versus local overlap between ME/CFS and psychiatric traits, and (iv) determining ME/CFS heritability cell-type enrichment.

## 2. Method

### 2.1 Ethics statement

The authors assert that all procedures contributing to this work comply with the ethical standards of the relevant national and institutional committees on human experimentation and with the Helsinki Declaration of 1975, as revised in 2013. This study is a secondary analysis of publicly available, de-identified GWAS summary statistics from previously published consortia; no individual-level data were accessed and no new data were collected. Ethical approval was therefore not required for this secondary analysis. Approvals and informed consent for the original studies were obtained by the respective consortia (DecodeME, FinnGen, the Psychiatric Genomics Consortium and UK Biobank), as described in the source publications.

### 2.2 Data sources

Five traits were included (Table 1). ME/CFS summary statistics for total-onset and infection-onset subgroups were from the DecodeME study^2^: 15,579 all-onset cases and 9,738 infection-onset cases (precipitating infection self-reported via validated questionnaire), each analysed against 259,909 UK Biobank controls (total N=275,488 and N=269,647, respectively). IBS summary statistics were from FinnGen Data Freeze 9 (K11_IBS; ICD-10 K58 hospital registry codes; 9,296 cases; 342,499 controls). ^8^ MDD statistics were from PGC MDD Wave 2 excluding 23andMe and UK Biobank (45,396 cases; 97,250 controls).^9^ Loneliness statistics were from the MRC IEU UK Biobank GWAS pipeline (ukb-b-8476; continuous; N=455,364).^10^ One trait intended as a fibromyalgia proxy was excluded following quality review (endpoint identity could not be verified).

**Table 1.** Study datasets.

| Trait | Source | Total N | Cases | Controls | $h^2_{\text{obs}}$ |
| --- | --- | --- | --- | --- | --- |
| ME/CFS (all cases) | DecodeME<sup>2</sup> | 275,488 | 15,579 | 259,909 | 0.016 |
| ME/CFS (infection-onset) | DecodeME<sup>2</sup> | 269,647 | 9,738 | 259,909 | 0.012 |
| IBS | FinnGen R9 K11_IBS<sup>8</sup> | 351,795 | 9,296 | 342,499 | 0.101 |
| MDD | PGC MDD2<sup>9</sup>† | 142,646 | 45,396 | 97,250 | 0.093 |
| Loneliness/isolation | UKB IEU ukb-b-8476<sup>10</sup> | 455,364 | — | — | 0.035 |

† PGC MDD Wave 2 excluding 23andMe and UK Biobank cohorts. $h^2_{\text{obs}}$ : observed-scale SNP heritability.

### 2.3 LDSC global genetic correlations

Summary statistics were processed using munge_sumstats.py (GenomicSEM^11^) against HapMap3. Genome-wide r_g_ and observed-scale h^2^ were estimated using bivariate LDSC^12^ with the European 1000 Genomes Phase 3 LD reference. Liability-scale h^2^ was not computed owing to uncertainty in ME/CFS and IBS population prevalence. Sample overlap was assessed via the cross-trait LDSC intercept matrix; there was minimal evidence of sample overlap (cross-trait intercept −0.002 for ME/CFS–IBS, consistent with no meaningful overlap). All r_g_ estimates with Bonferroni-corrected p<0.05 are reported.

### 2.4 Local genetic correlation analysis (LAVA)

Local genetic correlations were estimated using LAVA v0.1.5^7^ across 2,495 semi-independent European LD loci (hg19).^13^ Univariate local heritability was tested first (threshold p<0.05); bivariate tests were run for pairs where both traits passed. Sample overlap was modelled using the LDSC intercept matrix. Multiple testing was controlled using the Benjamini–Hochberg FDR procedure across all 4,232 bivariate tests performed in the five-trait analytic set; loci with FDR<0.05 are reported.

### 2.5 Colocalisation analysis

Colocalisation of ME/CFS-inf and IBS association signals at the chromosome 12q24.22 locus was assessed using coloc.abf v5.2^14^ with default priors (p1 = p2 = 10^−4^; p12 = 10^−5^). The full LAVA locus window (117,091,844–118,256,124 hg19) was used. After allele alignment (DecodeME A1/A2 to FinnGen alt/ref) and exclusion of palindromic SNPs (A/T or G/C with MAF > 0.4), 3,425 SNPs were retained. Five posterior probabilities were estimated: PP.H0 (no causal variant in either trait), PP.H1 (only ME/CFS-inf), PP.H2 (only IBS), PP.H3 (distinct causal variants), and PP.H4 (shared causal variant). Sensitivity analysis at p12 ∈ {10^−4^, 10^−5^, 10^−6^} is reported in supplementary materials.

### 2.6 MAGMA cell-type enrichment

Gene-level association statistics were computed using MAGMA v1.10^15^ with European 1000 Genomes LD. Cell-type enrichment used the Descartes Human single-cell atlas (77 cell types, Level 2),^16^ applying the gene-property mode (specificity quantiles, one-sided, direction=greater). Robustness was confirmed using a top-10% binary gene-set mode. FDR correction was applied within each trait.

### 2.7 Statistical power considerations

Statistical power was considered as follows. (i) LDSC: with observed-scale SNP heritabilities of 0.012– 0.101 and sample sizes from 142,646 (MDD) to 455,364 (loneliness), LDSC was better powered for moderate genome-wide genetic correlations than for local or colocalisation analyses;^12^ all reported r_g_ estimates substantially exceeded the level at which LDSC is typically informative. (ii) LAVA: based on published simulations,^7^ LAVA provides approximately 80% power to detect a local ρ of 0.5 when local heritability for both traits is ≥1×10^−4^ and the univariate filter (p<0.05) is met. For loci with very small local heritability, ρ is poorly identified and may converge to boundary values (±1.000); several FDR-significant ρ estimates reported here are boundary-estimated and should not be interpreted as perfect biological concordance. (iii) Colocalisation: adequate posterior probability for a shared causal variant (PP.H4>0.5) typically requires at least one trait to harbour a SNP with genome-wide significance (p<5×10^−8^) within the locus. No SNP reached this threshold at chromosome 12q24.22 (minimum p=2.2×10^−5^ for ME/CFS-inf; minimum p=8.2×10^−4^ for IBS); colocalisation did not identify a shared causal variant (PP.H4=0.007 at the default prior; PP.H1=0.548 indicates ME/CFS-inf carries regional signal at this locus that is not shared at the SNP level with IBS), as discussed in Section 3.2. (iv) MAGMA cell-type analysis: with ~18,000 protein-coding genes contributing to the gene-property regression, MAGMA was better suited to detect moderate cell-type enrichment in adequately powered traits than to detect small effects in lower-powered traits; the strong neural enrichment in ME/CFS (P=1.2×10^−7^) substantially exceeded the detection threshold, whereas the non-significance of immune cell-type enrichment should be interpreted in the context of both atlas-resolution limitations and reduced power for small effects.

### 2.8 Software and reproducibility

Analyses used LDSC v1.0.1, LAVA v0.1.5, coloc v5.2, MAGMA v1.10, and GenomicSEM (R package). Statistical analyses were performed in R version 4.3 (R Foundation for Statistical Computing, Vienna, Austria) and Python 3.10 on a Linux platform. All analysis code and derived intermediate output files (LAVA univariate and bivariate results, MAGMA gene-property output, coloc input/output, regional plot input) are deposited at the public GitHub repository https://github.com/gamie7/mecfs-ibs-cross-trait-analysis and archived at Zenodo under DOI 10.5281/zenodo.20596388.

## 3. Results

### 3.1 Global genetic correlations (Figure 1)

**Figure 1.**
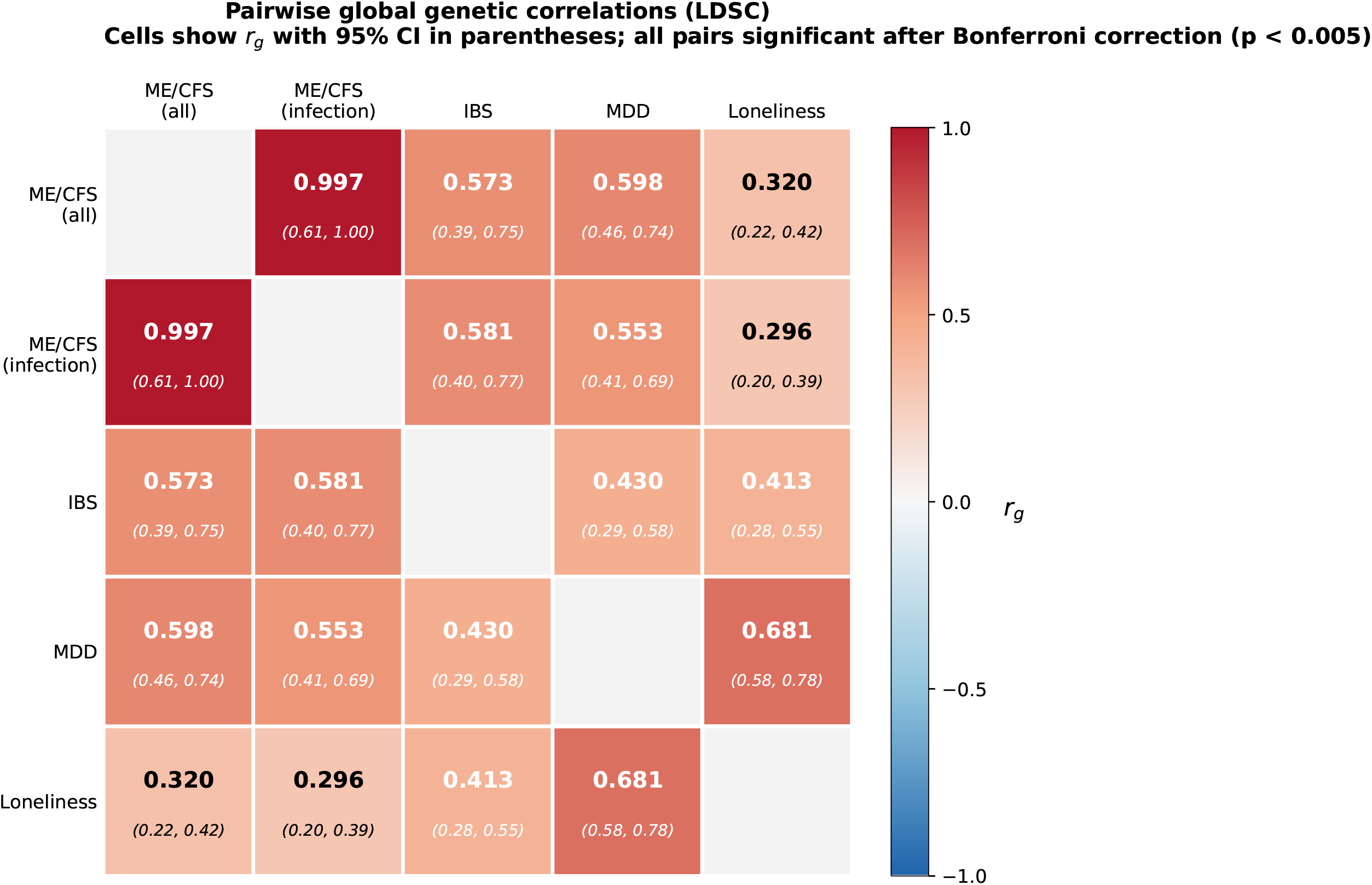
Global genetic correlations across five traits. Heatmap of pairwise global genetic correlations (r_g_) estimated using LDSC. Values are shown within cells; colour intensity reflects magnitude. All pairwise correlations were significant (p<0.05, Bonferroni-corrected). Diagonal cells are blank. h^2^: observed-scale SNP heritability. ME/CFS: myalgic encephalomyelitis/chronic fatigue syndrome; IBS: irritable bowel syndrome; MDD: major depressive disorder; inf: infection-onset subgroup.

All pairwise global r_g_ values were significant after Bonferroni correction (family-wise α=0.05 across 10 pairwise tests, corresponding to nominal p<0.005; Table 2; Figure 1). The highest values were between loneliness and MDD (r_g_=0.681), ME/CFS-all and MDD (r_g_=0.598) and ME/CFS-all and IBS (r_g_=0.573). Infection-onset ME/CFS showed slightly lower correlations with MDD (r_g_=0.553) and loneliness (r_g_=0.296) than all-cases ME/CFS, while showing a marginally higher r_g_ with IBS (0.581 vs 0.573). IBS showed moderate correlations with MDD (r_g_=0.430) and loneliness (r_g_=0.413). The near-perfect correlation between ME/CFS-all and ME/CFS infection-onset (r_g_=0.997) indicates that the infection-onset subgroup captures highly overlapping genetic architecture with the full case sample.^2^

**Table 2.**
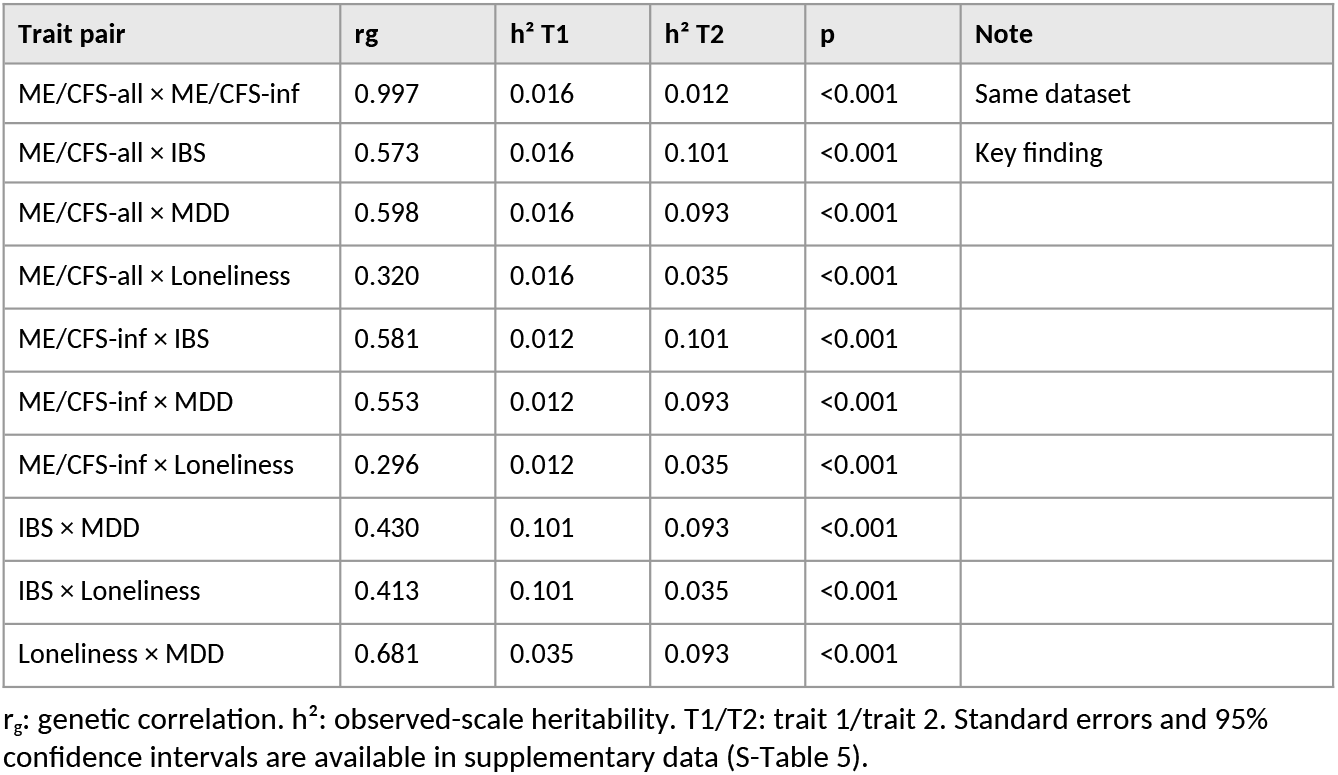
Pairwise global genetic correlations (LDSC). All Bonferroni-significant at family-wise α=0.05 (nominal p<0.005).

### 3.2 Local genetic correlations (Figure 2)

**Figure 2.**
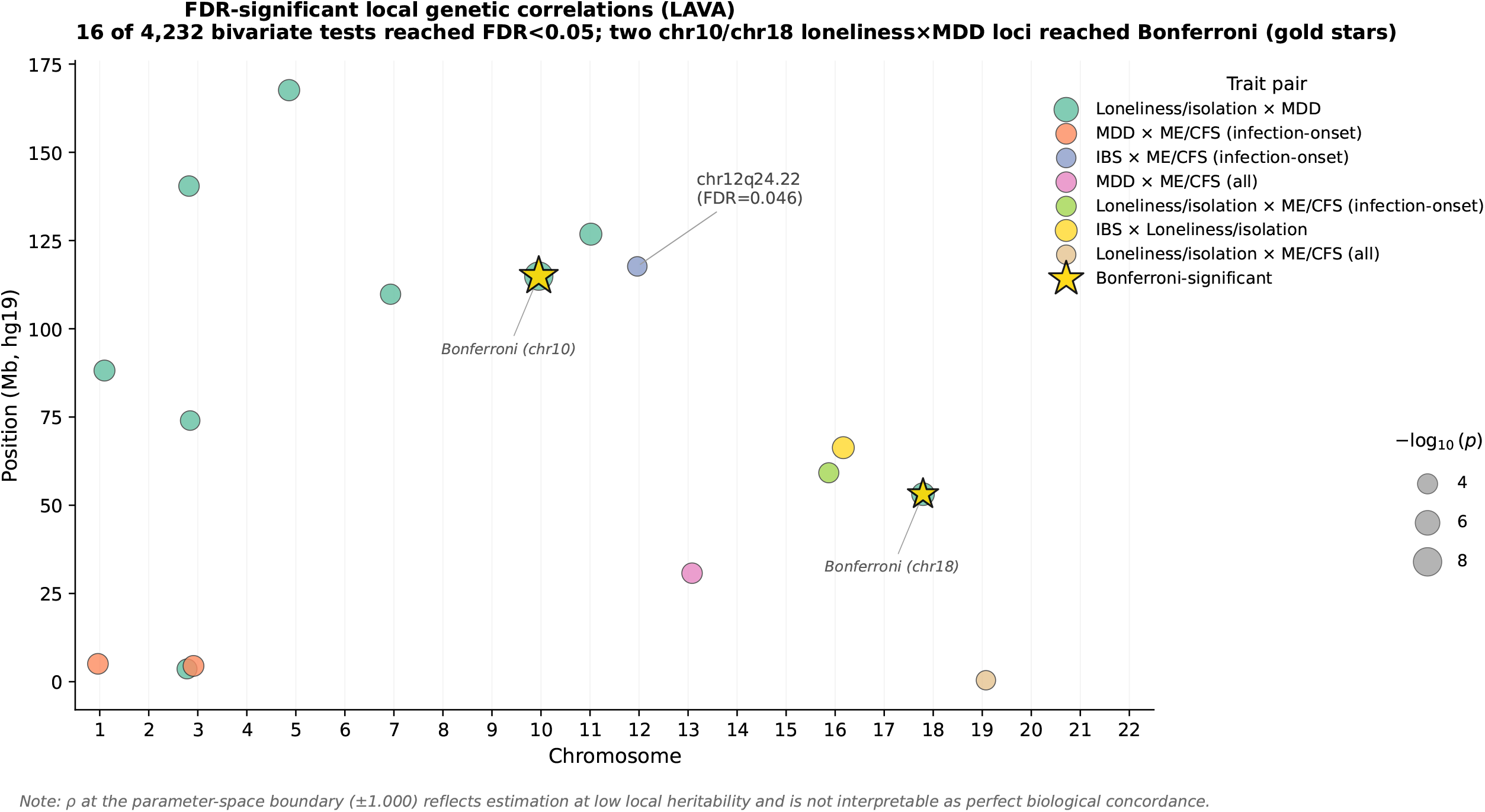
FDR-significant local genetic correlations (LAVA). Bubble plot of 16 FDR-significant loci across 4,232 bivariate tests. Each bubble represents one locus; x-axis: chromosome; y-axis: genomic position (Mb, hg19); bubble size: −log_10_(p). Colour indicates trait pair. Two loneliness×MDD loci (chr10, chr18) reached Bonferroni significance and are overlaid with gold stars. The chr12q24.22 (FDR=0.046) locus is annotated for orientation. ρ values at the parameter-space boundary (±1.000) reflect estimation at low local heritability and are not interpretable as perfect biological concordance.

Across 4,232 bivariate tests in nine trait pairs, 16 loci reached FDR<0.05 and two reached Bonferroni significance (both loneliness×MDD; Table 3; Figure 2). The loneliness×MDD pair showed the greatest number of FDR-significant loci (n=9), with the strongest signal at chromosome 10q23.33 (ρ=1.000; p=1.1×10^−8^; Bonferroni-significant). These Bonferroni-significant loneliness×MDD loci serve primarily as an internal benchmark, showing that the local-correlation framework identifies robust locus-level sharing for a closely related psychiatric pair, whereas ME/CFS–MDD local signals were FDR-only, boundary-estimated and not Bonferroni-robust.

**Table 3.** FDR-significant local genetic correlations (LAVA; 4,232 bivariate tests). *NOS1-encompassing locus.

| Trait 1 | Trait 2 | Chr | Position hg19 (bp) | Width (kb) | rho | p | FDR |
| --- | --- | --- | --- | --- | --- | --- | --- |
| Loneliness | MDD | 10 | 114,255,955–115,588,903 | 1,333 | 1.000 | $1.1 \times 10^{-8}$ | <0.001 |
| Loneliness | MDD | 18 | 52,512,524–53,762,996 | 1,250 | 0.750 | $8.0 \times 10^{-6}$ | 0.017 |
| Loneliness | MDD | 11 | 126,304,241–127,306,871 | 1,003 | 0.806 | $1.5 \times 10^{-5}$ | 0.017 |
| IBS | Loneliness | 16 | 65,882,821–66,738,843 | 856 | 1.000 | $1.6 \times 10^{-5}$ | 0.017 |
| Loneliness | MDD | 5 | 166,863,414–168,342,743 | 1,479 | 0.776 | $3.3 \times 10^{-5}$ | 0.028 |
| Loneliness | MDD | 1 | 87,667,573–88,656,689 | 989 | 0.892 | $4.3 \times 10^{-5}$ | 0.028 |
| ME/CFS-inf | MDD | 3 | 4,109,394–4,835,083 | 726 | 1.000 | $5.0 \times 10^{-5}$ | 0.028 |
| ME/CFS-inf | MDD | 1 | 4,773,404–5,362,404 | 589 | 0.969 | $5.7 \times 10^{-5}$ | 0.028 |
| Loneliness | MDD | 3 | 139,831,268–141,034,637 | 1,203 | –1.000 | $6.7 \times 10^{-5}$ | 0.028 |
| ME/CFS-all | MDD | 13 | 30,068,526–31,433,218 | 1,365 | 1.000 | $6.7 \times 10^{-5}$ | 0.028 |
| Loneliness | MDD | 7 | 109,095,559–110,479,479 | 1,384 | 0.745 | $8.2 \times 10^{-5}$ | 0.031 |
| ME/CFS-inf | Loneliness | 16 | 58,508,979–59,885,499 | 1,377 | 1.000 | $9.6 \times 10^{-5}$ | 0.034 |
| Loneliness | MDD | 3 | 3,128,153–4,109,393 | 981 | 1.000 | $1.1 \times 10^{-4}$ | 0.037 |
| Loneliness | MDD | 3 | 73,606,613–74,383,137 | 777 | 0.669 | $1.6 \times 10^{-4}$ | 0.046 |
| ME/CFS-inf | IBS* | 12 | 117,091,844–118,256,124 | 1,164 | 1.000 | $1.6 \times 10^{-4}$ | 0.046 |
| ME/CFS-all | Loneliness | 19 | 64,705–762,046 | 697 | –0.624 | $1.8 \times 10^{-4}$ | 0.049 |

Width: locus size in kilobases. p: local genetic correlation. Negative p: discordant genetic effects despite positive global $r_g$ . Two loci (chr10 and chr18, loneliness×MDD) reached Bonferroni significance ( $p < 1.2 \times 10^{-5}$ ). FDR computed across all 4,232 tests.

The primary ME/CFS-specific finding was a local genetic correlation between infection-onset ME/CFS and IBS at chromosome 12q24.22 (117,091,844–118,256,124 hg19; locus width 1,164 kb; ρ=1.000; p=1.6×10^−4^; FDR=0.046). This locus encompasses the *NOS1* gene (neuronal nitric oxide synthase; ENSG00000089250) and is spatially distinct from the DecodeME chr12q24.23 locus (*TAOK3*/*SUDS3*, ~118.5 Mb hg19), which lies in the adjacent LD block. This local genetic correlation is a borderline FDR result from a single locus. Colocalisation analysis (3,425 SNPs after allele alignment and palindromic SNP exclusion) did not identify a shared causal variant (PP.H4=0.007 at the default prior p12=10^−5^; PP.H1=0.548, PP.H0=0.351, PP.H3=0.057). The dominant PP.H1 indicates that ME/CFS-inf carries a regional polygenic signal at this locus that is not shared at the SNP level with IBS, where no SNP reached genome-wide significance (minimum p=2.2×10^−5^ for ME/CFS-inf; minimum p=8.2×10^−4^ for IBS). Within the locus, the strongest ME/CFS-inf SNPs cluster near the *TAOK3*-proximal edge (~118.2 Mb), whereas the strongest IBS SNPs cluster near *KSR2*/*NOS1* (~117.7 Mb); the regional plot in Figure S1 illustrates this separation. This pattern is consistent with the LAVA local correlation reflecting polygenic-level concordance across the window rather than a single shared causal variant. Sensitivity analysis at alternative priors (p12 ∈ {10^−4^, 10^−6^}) yielded PP.H4 between 0.0007 and 0.066, none supporting colocalisation (S-Table 9). The LAVA result captures correlated effect sizes across all SNPs in the locus and is not equivalent to confirmation of a shared causal variant. These findings should therefore be treated as hypothesis-generating, pending replication in larger samples. This signal nominally emerged in the infection-onset subgroup (ME/CFS-all×IBS: ρ=1.000; p=0.0020; FDR=0.118, not significant); given the near-identical genome-wide architecture of the two ME/CFS phenotypes (r_g_=0.997), the FDR difference at this locus may partly reflect the reduced power of the smaller infection-onset sample (n=9,738 vs n=15,579 all-cases) rather than a genuine biological subgroup distinction. Replication with larger infection-onset cohorts is needed.

ρ values at the parameter boundary (±1.000) occur in LAVA when local heritability is modest relative to the effective number of SNPs tested, and reflect estimation at the boundary of the permissible parameter space rather than perfect biological concordance between traits. This applies to several loci in Table 3 including the chromosome 12 *NOS1*-encompassing locus.

Infection-onset ME/CFS showed FDR-significant local correlations with MDD at two loci (chromosomes 1 and 3; ρ=0.969 and 1.000 respectively; both FDR=0.028) and with loneliness at one locus (chromosome 16; ρ=1.000; FDR=0.034); ME/CFS all-cases showed an FDR-significant local correlation with MDD at chromosome 13 (ρ=1.000; FDR=0.028). None of these ME/CFS–MDD or ME/CFS–loneliness signals reached Bonferroni significance, and all involved ρ estimates at or near the parameter-space boundary. Negative local genetic correlations were observed at chromosome 3 for loneliness×MDD (ρ=−1.000; FDR=0.028) and at chromosome 19 for ME/CFS-all×loneliness (ρ=−0.624; FDR=0.049), indicating loci with discordant local effects despite positive global genetic correlations.

Importantly, the chromosome 12q24.22 locus that showed an ME/CFS-IBS local correlation did not show a corresponding ME/CFS–MDD local correlation; ME/CFS–MDD local sharing therefore appears at different chromosomal positions to the ME/CFS–IBS locus rather than co-locating at it. The substantial global r_g_=0.598 between ME/CFS and MDD is accompanied by sparse, boundary-estimated and non-Bonferroni-significant local sharing rather than by a uniform locus-level architecture, consistent with DecodeME’s colocalisation findings showing no shared causal variants between ME/CFS and depression.^2^

### 3.3 ME/CFS cell-type enrichment (Figure 3)

**Figure 3.**
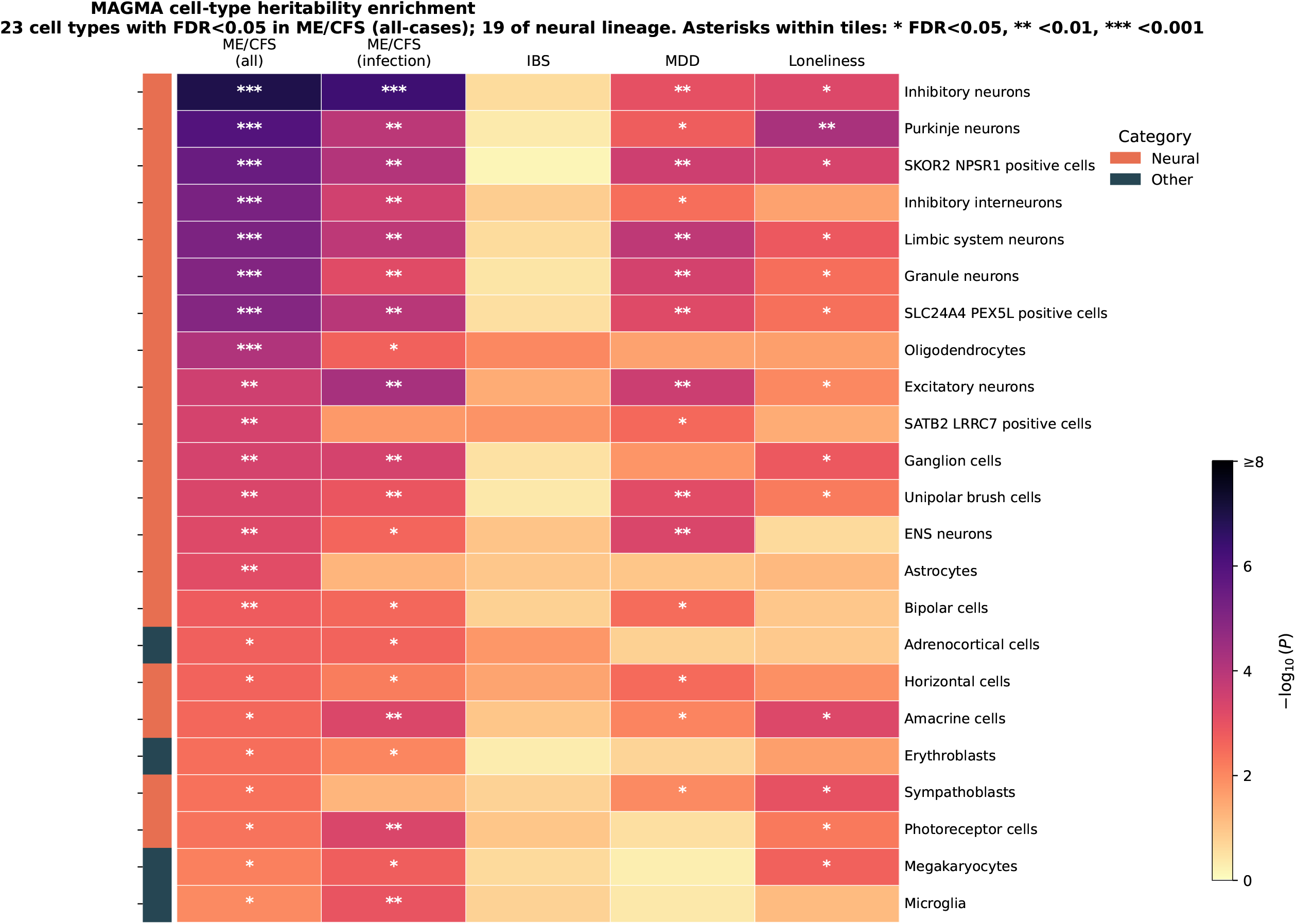
MAGMA cell-type enrichment in ME/CFS. Heatmap of MAGMA gene-property enrichment results (−log_10_ p) for FDR-significant cell types (FDR<0.05 in ME/CFS all-cases) across five traits. Annotation bar (left): orange = neural cell types; blue = other cell types. Columns: ME/CFS all-cases, ME/CFS infection-onset, IBS, MDD, loneliness. Descartes Human single-cell atlas, Level 2 (77 cell types). No peripheral immune cell types (B cells, T cells, antigen-presenting cells) reached FDR<0.05 in any ME/CFS phenotype; microglia (CNS-resident myeloid) reached FDR<0.05 in both ME/CFS phenotypes.

MAGMA gene-property analysis identified significant ME/CFS heritability enrichment in neural cell types (Figure 3). Inhibitory neurons showed the strongest signal (P=1.2×10^−7^; FDR<0.001), followed by Purkinje neurons, SKOR2/NPSR1-positive cells, inhibitory interneurons and limbic system neurons (all FDR<0.05; 19 of 23 FDR-significant cell types were of neural lineage, including 17 neurons and 2 glial populations). Infection-onset ME/CFS showed a near-identical enrichment profile. No peripheral immune cell types (B cells, T cells, antigen-presenting cells, peripheral myeloid populations) reached FDR significance in either ME/CFS phenotype; antigen-presenting cells reached nominal P<0.05 in ME/CFS-infection-onset (P=0.022) but did not survive multiple-testing correction (FDR=0.071). Microglia — the CNS-resident myeloid population — did reach FDR significance in both ME/CFS phenotypes (ME/CFS all-cases: P=0.011, FDR=0.036; ME/CFS infection-onset: P=0.0012, FDR=0.008), consistent with a neuroglial/neuroimmune component contributing to ME/CFS heritability rather than peripheral immune mechanisms specifically. This finding should be interpreted cautiously. The Descartes atlas is fetal-derived and may not adequately represent adult immune cell types, activated immune states or tissue-resident immune populations relevant to ME/CFS pathobiology. Absence of enrichment in this atlas does not exclude immune mechanisms. IBS showed no FDR-significant cell-type enrichment, likely reflecting limited power at the available sample size. Robustness was confirmed using a top-10% gene-set mode: Purkinje neurons, inhibitory interneurons and SATB2/LRRC7-positive cells reached FDR<0.05 in ME/CFS all-cases. Notably, the enteric nervous system neurons (ENS_neurons) category within the Descartes atlas showed FDR-significant heritability enrichment in both ME/CFS phenotypes (ME/CFS all-cases: P=6.2×10^−4^; FDR=0.004; ME/CFS infection-onset: P=2.6×10^−3^; FDR=0.012). This finding is biologically concordant with the *NOS1* locus signal (Section 3.2), since *NOS1* encodes the dominant enteric neuronal nitric oxide synthase, and it provides direct cell-type-level support for the involvement of enteric neural populations in ME/CFS heritability. IBS did not show FDR-significant ENS_neurons enrichment in this analysis (P=0.101), which may reflect the limited power of the current IBS sample for cell-type analyses. These findings are concordant with DecodeME’s brain tissue enrichment.^2^

## 4. Discussion

This study provides a systematic cross-trait genetic analysis of ME/CFS, IBS, and psychiatric traits. Three findings frame the contribution. First, ME/CFS–MDD genetic overlap was substantial at the global level (r_g_ ≈ 0.6) but only sparsely localised: of 4,232 bivariate local tests, three ME/CFS–MDD pairs reached FDR significance (chromosomes 1, 3 and 13), none reached Bonferroni significance, and all involved boundary-estimated ρ values close to 1.000 — a partial global–local dissociation rather than a complete absence of locus-level sharing. Second, ME/CFS heritability was strongly enriched in inhibitory neurons (P = 1.2 × 10^−7^) and, with biological coherence, in enteric nervous system neurons (FDR = 0.004), supporting neural and enteric neuronal contributions to disease susceptibility; the absence of peripheral immune cell-type enrichment in this atlas should not be interpreted as excluding immune mechanisms, and microglia (CNS-resident myeloid cells) did show FDR-significant enrichment in both ME/CFS phenotypes. Third, a borderline FDR-significant local correlation was observed between infection-onset ME/CFS and IBS at chromosome 12q24.22 — a genomic region containing *NOS1* — generating hypotheses regarding enteric neural mechanisms. We emphasise that the chromosome 12 finding is borderline (FDR = 0.046), that the LAVA ρ estimate sits at the parameter-space boundary, that colocalisation did not identify a shared causal variant (PP.H4 = 0.007), and that SNP-level peaks for the two traits fall in different sub-regions of the locus (Figure S1). It is therefore treated as polygenic-level evidence at a candidate locus, not as confirmation of a shared variant or specific gene.

Prior genetic work on ME/CFS comorbidity has been largely limited to epidemiological co-occurrence rates and genome-wide correlations. The present analysis adds resolution at both the locus level (LAVA across 2,495 LD blocks) and the cell-type level (MAGMA across 77 Descartes Human atlas Level-2 cell types), and, to our knowledge, is the first cross-trait LAVA analysis of the DecodeME GWAS, including its infection-onset subgroup.

### 4.1 Partial dissociation: ME/CFS–MDD global versus local overlap

ME/CFS–MDD overlap was characterised by substantial global genetic correlation (r_g_ = 0.598, 95% CI 0.46–0.74) paired with sparse and statistically uncertain local sharing. Three ME/CFS–MDD pairs reached FDR significance across 4,232 bivariate tests: ME/CFS infection-onset × MDD on chromosome 1 (ρ = 0.969; FDR = 0.028) and chromosome 3 (ρ = 1.000; FDR = 0.028), and ME/CFS all-cases × MDD on chromosome 13 (ρ = 1.000; FDR = 0.028). None of these reached Bonferroni significance, and all involved ρ estimates at or near the parameter-space boundary, which occurs when local heritability is modest relative to the effective number of SNPs and is not interpretable as perfect biological concordance. Importantly, the chromosome 12 locus that showed ME/CFS-IBS sharing did not show ME/CFS–MDD sharing. By comparison, the loneliness×MDD pair contributed 9 of the 16 FDR-significant local correlations and the only two Bonferroni-significant signals (chr10 and chr18), indicating that within this analytic framework MDD’s local genetic architecture is more sharply localisable with loneliness than with ME/CFS.

Two cautions are essential. Absence of Bonferroni-significant ME/CFS–MDD local signal does not exclude shared underlying mechanisms; the boundary-estimated ρ values reflect limited identification of local heritability for ME/CFS at current sample sizes rather than confirmation of distinct biology. The substantial global r_g_ between ME/CFS and MDD does suggest shared polygenic background — for example, neuroticism or distress-related traits. The pattern is best characterised as a partial global–local dissociation: locus-level sharing exists but is sparse, statistically uncertain and not robust to genome-wide multiple-testing correction. This is independently supported by DecodeME, whose colocalisation analysis did not detect shared causal variants between ME/CFS and depression.^2^ Taken together, the data do not support reducing ME/CFS to depression at the level of genetic architecture, while remaining consistent with the broader polygenic overlap reported in prior chronic fatigue genetic studies.

### 4.2 Neural and enteric cell-type enrichment

Neural cell-type enrichment of ME/CFS heritability, with FDR-significant microglial enrichment but without peripheral immune cell-type enrichment, is consistent with combined neural and CNS-resident myeloid contributions to ME/CFS susceptibility and replicates DecodeME brain tissue findings at single-cell resolution.^2^ Inhibitory neurons showed the strongest signal (P = 1.2 × 10^−7^), with broad enrichment across multiple neuronal subtypes (Purkinje neurons, granule neurons, limbic system neurons, excitatory neurons), supporting a CNS-level neurobiological component of susceptibility, while not excluding immune, autonomic or metabolic mechanisms that may be less well captured by the atlas and analytic design. ME/CFS heritability was additionally FDR-significantly enriched in enteric nervous system neurons (ME/CFS all-cases: P = 6.2 × 10^−4^; FDR = 0.004; ME/CFS infection-onset: FDR = 0.012). Enrichment of ENS neurons — despite the Descartes atlas being fetal-derived — indicates that enteric neural lineages are at least partially captured by the atlas and contribute meaningfully to ME/CFS heritability. The convergence of ME/CFS, IBS and fibromyalgia^21^ on neural enrichment patterns supports the hypothesis of a shared neurobiological substrate for functional somatic comorbidity. We note that the non-significance of peripheral immune cell-type enrichment should be interpreted in the context of both atlas-resolution limitations (the Descartes atlas may not adequately represent adult immune cell types or activated immune states) and reduced power for small effects; conversely, the FDR-significant microglial enrichment in both ME/CFS phenotypes is consistent with a neuroglial/neuroimmune component that warrants further investigation.

### 4.3 Hypothesis-generating evidence at the chromosome 12q24.22 locus

A borderline FDR-significant local genetic correlation between infection-onset ME/CFS and IBS was observed at chromosome 12q24.22 (FDR = 0.046), within a genomic region containing *NOS1. NOS1* encodes neuronal nitric oxide synthase, the dominant isoform regulating nitrergic neurotransmission in the enteric nervous system.^17^ Loss of NOS1 function in mice produces gut dysmotility,^18^ reduced nitrergic neuron density has been reported in human IBS,^19^ and autonomic and enteric nervous system dysfunction is increasingly recognised in ME/CFS.^20^ Concurrently, ME/CFS heritability was independently FDR-significantly enriched in enteric nervous system neurons (Section 4.2). Taken together, these observations generate hypotheses regarding enteric neural mechanisms in the ME/CFS–IBS overlap that warrant further investigation in larger post-infectious cohorts.

Several caveats apply. The local correlation is borderline (FDR = 0.046; a single locus surviving multiple-testing correction); the LAVA ρ estimate sits at the parameter-space boundary (1.000), which occurs when local heritability is modest relative to the effective number of SNPs and should not be read as perfect biological concordance; and colocalisation analysis explicitly did not identify a single shared causal variant (PP.H4 = 0.007 at the default prior). Within the locus, the strongest ME/CFS-inf SNP-level signals cluster near 118.2 Mb (annotated to *TAOK3* by FinnGen, although this region also falls within the *KSR2* gene body), whereas the strongest IBS signals cluster near 117.7–117.8 Mb (within *KSR2*, near *NOS1*); the regional plot in Figure S1 illustrates this separation. The pattern is consistent with the LAVA correlation reflecting polygenic-level concordance across the window rather than a single shared causal variant. LAVA detects correlated polygenic effects across a locus window and does not identify causal variants, genes, or mechanisms; “the *NOS1* locus” should be read as a description of the genomic window rather than as identification of *NOS1* itself as the responsible gene. The biological interpretation of this signal must await both independent replication in larger post-infectious cohorts and functional follow-up.

### 4.4 Infection-onset specificity

The chromosome 12 signal emerged in the infection-onset ME/CFS subgroup rather than in the all-cases group, despite the two phenotypes sharing near-identical genome-wide architecture (r_g_ = 0.997). The FDR difference at this locus (ME/CFS-inf×IBS: FDR = 0.046; ME/CFS-all×IBS: FDR = 0.118) may reflect reduced phenotypic heterogeneity in the infection-onset subgroup, although this interpretation is uncertain because the subgroup is smaller and the genome-wide architecture is nearly identical to all-cases ME/CFS. Post-infectious IBS is similarly thought to involve distinct neurobiological mechanisms. ^4^ These observations support further genetic studies of infection-onset phenotypes, including in larger post-COVID-19 cohorts where such stratification may become routinely feasible.

### 4.5 Clinical implications

Three implications follow, each appropriately hedged by the hypothesis-generating nature of the analysis. First, the partial global–local dissociation between ME/CFS and MDD provides genetic evidence that does not support reducing ME/CFS to depression at the level of genetic architecture: substantial polygenic overlap coexists with sparse, boundary-estimated and non-Bonferroni-robust local sharing, with no shared signal at the chromosome 12 ME/CFS-IBS locus. Second, if the chromosome 12q24.22 signal is replicated in larger cohorts, a locus containing *NOS1* would provide a candidate region connecting post-infectious ME/CFS and IBS at the enteric neuromotor level, an area accessible to pharmacological exploration. Third, infection-onset subgroup stratification, where feasible, may help identify more biologically homogeneous groups in genetic studies of post-infectious multimorbidity — a category that has become more clinically prominent following SARS-CoV-2.

### 4.6 Limitations

The following limitations should be considered (statistical power for each method is detailed in Section 2.7). First, the Descartes Human atlas is fetal-derived and may not fully capture adult-specific enteric neuron subtypes or activated immune states; although ENS_neuron populations within the atlas did show FDR-significant ME/CFS heritability enrichment (Section 3.3), more comprehensive adult gut atlases (e.g. the Gut Cell Atlas)^22^ may reveal additional cell-type-specific signals and merit follow-up analysis. Second, the chromosome 12 *NOS1*-encompassing locus finding is a borderline FDR result (FDR=0.046) from a single locus; colocalisation did not identify a shared causal variant (PP.H4=0.007) and indicated distinct SNP-level peaks for the two traits within the locus, consistent with polygenic-level correlation rather than a single shared variant. The finding is hypothesis-generating and requires independent replication. Third, IBS in FinnGen is defined by hospital discharge registry codes (ICD-10 K58), likely capturing more severe or clinically diagnosed IBS and potentially under-representing community presentations. Fourth, this is a secondary analysis and causal inference is not possible. Fifth, observed-scale heritabilities are reported; liability-scale estimates depend on assumed population prevalences for ME/CFS and IBS, which remain uncertain. Sixth, ME/CFS infection-onset was defined by self-report, and the generalisability of findings to objectively confirmed post-infectious ME/CFS requires evaluation. Seventh, no independent GWAS cohort replication of the chromosome 12 locus was performed.

### 4.7 Summary

In summary, this cross-trait analysis identifies a partial dissociation between global and local genetic architecture in the ME/CFS–MDD relationship: a substantial global genetic correlation (r_g_ = 0.598) is accompanied by three FDR-significant but non-Bonferroni-significant, boundary-estimated local correlations at chromosomes 1, 3 and 13, with no ME/CFS–MDD sharing at the chromosome 12 ME/CFS–IBS locus. The pattern does not support reducing ME/CFS to depression at the level of genetic architecture, while remaining consistent with broader polygenic overlap. ME/CFS heritability is strongly enriched in inhibitory neurons (P = 1.2 × 10^−7^) and, with biological coherence, in enteric nervous system neurons (FDR = 0.004), supporting neural and enteric neuronal contributions to disease susceptibility; the absence of peripheral immune cell-type enrichment in this atlas should not be interpreted as excluding immune mechanisms, and microglia did show FDR-significant enrichment in both ME/CFS phenotypes. A borderline FDR-significant local correlation between infection-onset ME/CFS and IBS at chromosome 12q24.22 — a region containing *NOS1* — generates hypotheses regarding enteric neural mechanisms; colocalisation did not identify a shared causal variant, and this signal is offered as a candidate for further investigation rather than as established. Replication in larger post-infectious cohorts, functional study of enteric nitrergic pathways, and continued differentiation of ME/CFS from depression in clinical practice are warranted.

## Supporting information

STREGA Checklist

## Author contributions

J.H.L. conceived the study, designed and performed all analyses, interpreted the results, wrote and revised the manuscript, and approved the final version for publication. The author is accountable for all aspects of the work.

## Transparency declaration

The lead author affirms that this manuscript is an honest, accurate and transparent account of the study being reported, that no important aspects of the study have been omitted, and that any discrepancies from the study as planned have been explained.

## Data availability

All summary statistics analysed in this study are publicly available from the original consortia: DecodeME (doi:10.1101/2025.08.06.25333109); FinnGen R9 (https://storage.googleapis.com/finngen-public-data-r9/); PGC MDD (https://pgc.unc.edu); UK Biobank loneliness/isolation (https://gwas.mrcieu.ac.uk; ukb-b-8476). Full GWAS-level summary statistics including p-values, effect directions and standard errors for all tested markers are available from the original consortia as listed. All analysis code and derived intermediate output files are available at the public GitHub repository https://github.com/gamie7/mecfs-ibs-cross-trait-analysis, archived at Zenodo under DOI 10.5281/zenodo.20596388. The repository includes the LAVA univariate and bivariate results, MAGMA gene-property output, coloc input/output and regional plot input, plus an annotated reproducibility document specifying software versions and execution order.

## Acknowledgements

The author thanks the participants and investigators of the DecodeME study, the FinnGen consortium, the Psychiatric Genomics Consortium and UK Biobank, whose publicly shared summary statistics made this work possible.

## Artificial intelligence use

The author used Claude (Anthropic) via the claude.ai interface for code-drafting support during preparation of R/Python analysis scripts. The tool was not used to generate or modify numerical results, select statistical methods, interpret findings, conduct literature reviews, or write substantive scientific conclusions. All code was reviewed, edited and executed by the author, and all numerical outputs were independently checked against the original analysis files. The author takes full responsibility for the manuscript, analyses, interpretation, references and conclusions.

## Figure legends

**Figure S1 (supplementary). Regional association plot at chr12q24.22**. Three-panel figure showing single-SNP association −log_10_(p) values across the LAVA locus (117.0–118.3 Mb hg19) for (A) infection-onset ME/CFS (DecodeME; minimum p = 2.2×10^−5^) and (B) IBS (FinnGen R9 K11_IBS; minimum p = 8.2×10^−4^), with SNPs coloured by nearest annotated gene; (C) protein-coding gene track for the locus. Genome-wide significance (p = 5×10^−8^) and suggestive (p = 10^−5^) thresholds are shown as dashed and dotted lines respectively. The strongest signals for the two traits cluster in different sub-regions of the locus; coloc PP.H4 = 0.007 at the default prior, with no shared causal variant identified. The *TAOK3* gene extends past the locus boundary and is shown for orientation.

## References

1. Lim EJ, Ahn YC, Jang ES, Lee SW, Lee SH, Son CG. Systematic review and meta-analysis of the prevalence of chronic fatigue syndrome/myalgic encephalomyelitis. J Transl Med. 2020;18:100.

2. DecodeME Collaboration. Initial findings from the DecodeME genome-wide association study of myalgic encephalomyelitis/chronic fatigue syndrome. medRxiv. 2025. doi:10.1101/2025.08.06.25333109.

3. Hickie I, Davenport T, Wakefield D, Vollmer-Conna U, Cameron B, Vernon SD, et al. Post-infective and chronic fatigue syndromes precipitated by viral and non-viral pathogens: prospective cohort study. BMJ. 2006;333:575.

4. Spiller R, Garsed K. Postinfectious irritable bowel syndrome. Gastroenterology. 2009;136:1979–1988.

5. Aaron LA, Buchwald D. A review of the evidence for overlap among unexplained clinical conditions. Ann Intern Med. 2001;134:868–881.

6. Eijsbouts C, Zheng T, Kennedy NA, Bonfiglio F, Anderson CA, Moutsianas L, et al. Genome-wide analysis of 53,400 people with irritable bowel syndrome highlights shared genetic pathways with mood and anxiety disorders. Nat Genet. 2021;53:1543–1552.

7. Werme J, van der Sluis S, Posthuma D, de Leeuw CA. An integrated framework for local genetic correlation analysis. Nat Genet. 2022;54:274–282.

8. Kurki MI, Karjalainen J, Palta P, Sipilä TP, Kristiansson K, Donner KM, et al. FinnGen provides genetic insights from a well-phenotyped isolated population. Nature. 2023;613:508–518.

9. Wray NR, Ripke S, Mattheisen M, Trzaskowski M, Byrne EM, Abdellaoui A, et al. Genome-wide association analyses identify 44 risk variants and refine the genetic architecture of major depression. Nat Genet. 2018;50:668–681.

10. Elsworth B, Lyon M, Alexander T, Liu Y, Matthews P, Hallett J, et al. The MRC IEU OpenGWAS data infrastructure. bioRxiv. 2020. doi:10.1101/2020.08.10.244293.

11. Grotzinger AD, Rhemtulla M, de Vlaming R, Ritchie SJ, Mallard TT, Hill WD, et al. Genomic structural equation modelling provides insights into the multivariate genetic architecture of complex traits. Nat Hum Behav. 2019;3:513–525.

12. Bulik-Sullivan BK, Loh PR, Finucane HK, Ripke S, Yang J, Patterson N, et al. LD Score regression distinguishes confounding from polygenicity in genome-wide association studies. Nat Genet. 2015;47:291–295.

13. Berisa T, Pickrell JK. Approximately independent linkage disequilibrium blocks in human populations. Bioinformatics. 2016;32:283–285.

14. Giambartolomei C, Vukcevic D, Schadt EE, Franke L, Hingorani AD, Wallace C, et al. Bayesian test for colocalisation between pairs of genetic association studies using summary statistics. PLoS Genet. 2014;10:e1004383.

15. de Leeuw CA, Mooij JM, Heskes T, Posthuma D. MAGMA: generalised gene-set analysis of GWAS data. PLoS Comput Biol. 2015;11:e1004219.

16. Domcke S, Hill AJ, Daza RM, Fan J, O’Day DR, Pliner H, et al. A human cell atlas of fetal chromatin accessibility. Science. 2020;370:eaba7612.

17. Furness JB. The enteric nervous system and neurogastroenterology. Nat Rev Gastroenterol Hepatol. 2012;9:286–294.

18. Huang PL, Dawson TM, Bredt DS, Snyder SH, Fishman MC. Targeted disruption of the neuronal nitric oxide synthase gene. Cell. 1993;75:1273–1286.

19. Takahashi T. Pathophysiological significance of neuronal nitric oxide synthase in the gastrointestinal tract. J Gastroenterol. 2003;38:421–430.

20. Maksoud R, du Preez S, Eaton-Fitch N, Thapaliya K, Barnden L, Cabanas H, et al. A systematic review of neurological impairments in myalgic encephalomyelitis/chronic fatigue syndrome using neuroimaging techniques. PLoS One. 2020;15:e0232475.

21. Kerrebijn I, Bjornsdottir G, Arbabi K, Urpa L, Haapaniemi H, Thorleifsson G, et al. The genetic architecture of fibromyalgia across 2.5 million individuals. medRxiv. 2025. doi:10.1101/2025.09.18.25335914.

22. Elmentaite R, Kumasaka N, Roberts K, Fleming A, Dann E, King HW, et al. Cells of the human intestinal tract mapped across space and time. Nature. 2021;597:250–255.

