## Supplementary Material for "Global and local genetic overlap among ME/CFS, irritable bowel syndrome and psychiatric traits: a hypothesis-generating analysis"

### Supplementary Materials

*Cross-trait local genetic correlation of infection-onset ME/CFS, irritable bowel syndrome, and psychiatric traits: a hypothesis-generating analysis*

Jun Hyun Lee

#### Contents

S1. Description of supplementary tables (companion file: Supplementary_Tables.xlsx)

S2. Extended methods

S3. Supplementary table legends

S4. Reporting checklist note (STREGA)

S5. Detailed AI tool use disclosure

#### S1. Description of supplementary tables

All supplementary tables are provided in the companion workbook **Supplementary_Tables.xlsx**, which contains a **README** sheet with a complete column dictionary, highlight legend and reproducibility checks. The ten tables included are:

- **S-Table 1 (LAVA univariate).** Local SNP heritability ($h^2$) for each of 6 traits across 2,495 Berisa–Pickrell LD blocks (hg19). 11,520 rows. Includes the excluded MCP proxy for transparency.
- **S-Table 2 (LAVA bivariate, full).** All bivariate local genetic correlations across 14 trait pairs (includes MCP). 6,714 rows. No multiple-testing columns — applied only to the analytic set in S-Table 3.
- **S-Table 3 (LAVA bivariate, analytic — PRIMARY).** 4,232 bivariate tests across 9 trait pairs after MCP exclusion, with Benjamini–Hochberg FDR and Bonferroni adjustments. 16 loci FDR-significant; the chr12q24.22 *NOS1*-encompassing locus (locus_i 1846) highlighted in green.
- **S-Table 4 (MAGMA top-10% sensitivity).** Top-10% binary gene-set robustness analysis: 77 Descartes Human atlas Level-2 cell types × 5 traits = 385 tests.
- **S-Table 5 (LDSC global rg).** Global genetic correlations across 15 pairs from 6 traits. rg, SE, 95% CI, Z, p, intercept terms. Point estimates reproduce main-text Table 2 to 3 decimal places.
- **S-Table 6 (LDSC intercept matrix).** 6×6 matrix: $h^2$ intercept on diagonal, cross-trait intercept off-diagonal. Used to model sample overlap in LAVA.
- **S-Table 7 (MAGMA gene-property primary — PRIMARY behind Figure 3).** Continuous specificity-quantile mode (TYPE=COVAR), condition-hide=Average, direction=greater. 77 cell types × 5 traits. Within-trait FDR.
- **S-Table 8 (Coloc input SNPs).** 3,425 SNPs at the chr12q24.22 LAVA locus (117,091,844–118,256,124 hg19) after allele alignment and palindromic SNP exclusion (A/T or G/C with MAF >

0.4). NOS1 gene-body SNPs highlighted green; SNPs with $p<5\times10^{-5}$ in either trait highlighted yellow.

- **S-Table 9 (Coloc sensitivity).** Posterior probabilities (PP.H0–H4) at three prior settings (p12 = $10^{-4}$, $10^{-5}$, $10^{-6}$). Default prior row highlighted.
- **S-Table 10 (Locus genes).** Protein-coding genes (n=8) and all biotypes (n=17) within the chr12q24.22 locus, queried from Ensembl GRCh37. *NOS1* highlighted green.

#### S2. Extended methods

##### S2.1 Summary-statistics quality control and harmonisation

All five primary summary-statistics files were processed with `munge_sumstats.py` from the LDSC distribution (GenomicSEM wrapper). HapMap3 SNP filter was applied. MAF threshold was 0.01 and INFO threshold (where available) was 0.9. Effect allele and reference allele were aligned to the HapMap3 reference; ambiguous strand-flip palindromic SNPs (A/T, G/C) with MAF>0.4 were removed before LDSC and LAVA analyses.

##### S2.2 LDSC global genetic correlations

Bivariate LDSC was run with the European 1000 Genomes Phase 3 LD reference via the GenomicSEM `ldsc()` wrapper. Genome-wide $r_g$ was extracted from the genetic-covariance matrix S as $S[i,j]/\sqrt{(S[i,i]\cdot S[j,j])}$; standard errors were computed via the delta method using diagonal elements of the V matrix (saved as S-Table 5). For highly overlapping datasets (e.g. ME/CFS-all × ME/CFS-inf), the diagonal-approximation SE may be conservative; original LDSC.py block-jackknife SEs would be smaller. The cross-trait LDSC intercept matrix (S-Table 6) was used to model sample overlap in the LAVA bivariate step. The ME/CFS × IBS cross-trait intercept (~ −0.003) was consistent with no meaningful overlap, as DecodeME uses UK Biobank controls and FinnGen R9 uses a non-overlapping Finnish cohort.

##### S2.3 LAVA univariate and bivariate testing

LAVA v0.1.5 was used with the 2,495 Berisa–Pickrell LD blocks (European; hg19). The univariate filter ($p<0.05$ for local $h^2$) gates inclusion in the bivariate step, as recommended by the LAVA authors. Bivariate testing was performed for each pair of filter-passing traits at each locus, with sample overlap modelled via the LDSC cross-trait intercept matrix supplied through the `sample.overlap` argument. The resulting 4,232 bivariate tests in the five-trait analytic set constitute the basis for multiple-testing correction (Benjamini–Hochberg FDR and Bonferroni; S-Table 3).

$\rho$ estimates at the parameter boundary (±1.000) occur when the locus is small relative to the effective number of SNPs tested; $\rho$ is bounded by the parameter space and can converge to the boundary. The chr12q24.22 $\rho=1.000$ is not interpreted as perfect concordance but as evidence of non-zero local correlation; the 95% confidence interval bounds (also in S-Table 3) are more informative.

##### S2.4 Colocalisation analysis

Colocalisation at the chr12q24.22 locus was performed using `coloc.abf()` from the `coloc` R package v5.2 with default priors: p1 = p2 = $10^{-4}$ (prior probability that a SNP is causal for one trait) and p12 = $10^{-5}$ (prior probability of shared causality). Input data were the full LAVA locus window (117,091,844–118,256,124 hg19; 1,164 kb) drawn from the original DecodeME ME/CFS infection-onset GWAS (rsID-mapped) and FinnGen R9 K11_IBS GWAS. After restriction to the locus, merging by rsID, allele alignment (DecodeME A1/A2 to FinnGen alt/ref; flipped-allele beta signs reversed, MAF inverted accordingly), and exclusion of palindromic SNPs (A/T or G/C with MAF > 0.4), **3,425 SNPs** were retained. Five posterior probabilities were estimated: PP.H0 (no causal variant in either trait), PP.H1 (only ME/CFS-inf), PP.H2 (only IBS), PP.H3 (distinct causal variants), and PP.H4 (shared causal variant).

**Result at default prior (p12 = $10^{-5}$):** PP.H0=0.351; **PP.H1=0.548**; PP.H2=0.037; PP.H3=0.057; **PP.H4=0.007**. The dominant PP.H1 indicates that ME/CFS-inf carries a regional polygenic signal at this locus that is not shared at the SNP level with IBS. Minimum p-values within the locus were $2.2\times10^{-5}$ for ME/CFS-inf and $8.2\times10^{-4}$ for IBS, neither reaching genome-wide significance. The strongest ME/CFS-inf SNPs cluster near the *TAOK3*-proximal edge of the locus (~118.2 Mb), whereas the strongest IBS SNPs cluster near *KSR2*/*NOS1* (~117.7 Mb). The full input SNP set is reported in S-Table 8.

**Sensitivity analysis.** PP.H4 was robust across prior settings: $6.6\times10^{-2}$ at the permissive prior (p12=$10^{-4}$), $7.1\times10^{-3}$ at default (p12=$10^{-5}$), and $7.1\times10^{-4}$ at the conservative prior (p12=$10^{-6}$). None supports colocalisation. Full sensitivity output is in S-Table 9.

#### S2.5 MAGMA cell-type enrichment

MAGMA v1.10 was used with European 1000 Genomes LD. The gene window was the default 0 kb up / 0 kb down (gene-body only) for the primary analysis. Cell-type specificity scores were derived from the Descartes Human single-cell atlas Level 2 (Domcke et al. 2020; 77 cell types). Two complementary analyses were performed:

- **Primary — gene-property (continuous specificity quantile) mode (S-Table 7).** One-sided test, direction = greater, conditioning on the across-cell-type average expression. Within each trait, p-values across 77 cell types were FDR-corrected. The Inhibitory_neurons signal in ME/CFS-all (P=$1.22\times10^{-7}$) reproduces the main-text value (P=$1.2\times10^{-7}$). Among non-neuronal lineages, microglia (CNS-resident myeloid) reached FDR<0.05 in both ME/CFS phenotypes (ME/CFS all-cases: P=0.011, FDR=0.036; infection-onset: P=0.0012, FDR=0.008), whereas no peripheral immune cell types (B cells, T cells, antigen-presenting cells, peripheral myeloid populations) reached FDR significance.
- **Sensitivity — top-10% binary gene-set mode (S-Table 4).** The top decile of genes by specificity score for each cell type was defined as a gene set and tested for enrichment against each trait. Three cell types (Purkinje neurons, inhibitory interneurons, SATB2/LRRC7-positive cells) reached FDR<0.05 in ME/CFS all-cases. As in the primary analysis, no peripheral immune cell types reached FDR significance.

#### S2.6 Software versions and computational environment

Analyses were performed on a Linux platform using: R 4.3.x; Python 3.10.x; LDSC v1.0.1

(https://github.com/bulik/ldsc); LAVA v0.1.5 (`github.com/josefin-werme/LAVA`); coloc R package v5.2; MAGMA v1.10; GenomicSEM R package (latest at time of analysis). All analysis code and derived intermediate output files are available at the public GitHub repository https://github.com/gamie7/mecfs-ibs-cross-trait-analysis, archived at Zenodo under DOI 10.5281/zenodo.20596388. See docs/reproducibility.md in the repository for the full software-version manifest and execution order.

##### S2.7 Excluded analysis: MCP proxy

One trait initially included as a fibromyalgia proxy was removed following pre-analysis quality review. The endpoint corresponds to a multi-site chronic pain (MCP) phenotype derived from UK Biobank questionnaire items rather than a clinically confirmed fibromyalgia diagnosis; the identity of the underlying phenotype against the fibromyalgia construct of interest could not be verified to the standard required for inclusion as a primary trait. The MCP results are retained in S-Tables 1, 2, 5 and 6 for full transparency about the exclusion; all primary multiple-testing correction and all main-text findings are based exclusively on the post-exclusion 5-trait, 4,232-test analytic set.

#### S3. Supplementary figure and table legends

**Figure S1. Regional association plot at chr12q24.22.** Three-panel figure illustrating the colocalisation analysis underlying Section 2.4. **Panel A:** infection-onset ME/CFS (DecodeME) single-SNP $-\log_{10}(p)$ values across the LAVA locus (117.0–118.3 Mb hg19); minimum $p = 2.2\times10^{-5}$. **Panel B:** IBS (FinnGen R9 K11_IBS) single-SNP $-\log_{10}(p)$ values across the same window; minimum $p = 8.2\times10^{-4}$. **Panel C:** protein-coding gene track for the locus, showing *C12orf49*, *RNFT2*, *HRK*, *FBXW8*, *TESC*, *FBXO21*, *NOS1*, *KSR2* (within locus) and the proximal end of *TAOK3* (extending past the locus boundary). SNPs are coloured by nearest annotated gene (FinnGen annotation). Genome-wide significance ($p = 5\times10^{-8}$) and suggestive ($p = 10^{-5}$) thresholds shown as red dashed and grey dotted lines respectively. The strongest signals for the two traits cluster in different sub-regions of the locus; colocalisation analysis (S-Table 9) did not identify a shared causal variant under any prior setting (PP.H4 ≤ 0.066). Figure source data: S-Table 8 (3,425 SNPs after harmonisation) and S-Table 10 (gene coordinates).

**S-Table 1. LAVA univariate local heritability.** Local SNP heritability ($h^2$) for each of six traits (ME/CFS all-cases, ME/CFS infection-onset, IBS, MDD, loneliness/isolation, and the excluded MCP proxy) at each of 2,495 Berisa–Pickrell LD blocks (hg19). 11,520 rows. Loci with univariate $p<0.05$ were carried forward to bivariate testing.

**S-Table 2. LAVA bivariate — full unfiltered set.** Bivariate ρ for all trait pairs at all loci passing the univariate filter, including pairs with the excluded MCP proxy (6,714 rows). No multiple-testing columns.

**S-Table 3. LAVA bivariate — primary analytic set.** 4,232-test analytic set after MCP exclusion. Columns as in S-Table 2 plus FDR (Benjamini–Hochberg) and BONF (Bonferroni). 16 loci FDR-significant; 2 loci (chr10 and chr18, loneliness×MDD) Bonferroni-significant. Yellow highlight = FDR<0.05; green =

chr12q24.22 *NOS1*-encompassing locus.

**S-Table 4. MAGMA top-10% binary gene-set sensitivity.** Robustness analysis: 77 Descartes Human atlas Level-2 cell types × 5 traits = 385 tests.

**S-Table 5. LDSC global rg.** Global genetic correlations across 15 trait pairs from 6 traits. Columns: rg, rg.SE, 95% CI, Z, p, $h^2$ for each trait, $h^2$-intercept for each trait, cross-trait intercept. SE/CI from delta method (diagonal V).

**S-Table 6. LDSC intercept matrix (6×6).** $h^2$-intercept on the diagonal (shaded blue); cross-trait intercept off-diagonal. Diagonal values near 1.0 indicate well-calibrated GWAS; off-diagonal values near 0 indicate independent samples.

**S-Table 7. MAGMA gene-property primary analysis.** Primary cell-type enrichment behind Figure 3. 5 traits × 77 Descartes Human atlas Level-2 cell types. Continuous specificity-quantile mode, TYPE=COVAR. Within-trait FDR.

**S-Table 8. Coloc input SNPs.** 3,425 SNPs at chr12q24.22 (117,091,844–118,256,124 hg19) after harmonisation. Both betas oriented to DecodeME A1 = effect allele. Columns include nearest gene annotation, MAF, beta+SE+p for each trait. Green highlight = SNPs within the *NOS1* gene body; yellow highlight = nominal significance ($p<5\times10^{-5}$) in either trait.

**S-Table 9. Coloc sensitivity.** PP.H0–H4 at three prior settings: p12 = $10^{-4}$ (permissive), $10^{-5}$ (default), $10^{-6}$ (conservative). Default-prior row highlighted orange. PP.H4 < 0.07 across all settings; no support for colocalisation.

**S-Table 10. chr12q24.22 locus genes.** Protein-coding genes (8 entries) and all biotypes (17 entries, including lincRNAs and pseudogenes) within the 1,164-kb locus, from Ensembl GRCh37 via biomaRt. *NOS1* (ENSG00000089250) at 117,645,947–117,889,975 highlighted green.

#### S4. Reporting checklist note

This study is a secondary cross-trait genetic correlation analysis of publicly available GWAS summary statistics. The most appropriate reporting framework is the STREGA (STrengthening the REporting of Genetic Association studies) extension to STROBE. A completed STREGA checklist will accompany the submission as a separate file; key items are addressed as follows:

- Title and abstract: structured abstract with effect sizes reported in Tables 2–3 and S-Tables 3, 5 (Items 1a–b).
- Study design: secondary analysis of summary statistics; design and rationale described in Introduction and Methods (Items 4–6).
- Participants: sourced from DecodeME, FinnGen R9, PGC MDD Wave 2, and UK Biobank; described in main-text Section 2.2 and Table 1 (Item 7).
- Variables: case/control definitions and continuous-trait scaling described per dataset (Item 7).

- Quantitative variables and statistical methods: methods described in main-text Section 2 and elaborated in Section S2 of these supplementary materials (Items 11–13).
- Bias: sample overlap modelled via LDSC cross-trait intercept (S-Table 6); ICD-K58 phenotype caveat noted in Limitations (Items 9–10).
- Study size and power: detailed in main-text Section 2.7 (Item 13).
- Statistical methods including handling of missing data and multiple testing: Section S2 of these materials (Items 12, 16).
- Results: descriptive and main results in main-text Sections 3.1–3.3; effect sizes with 95% CIs in Tables 2–3 and S-Tables 3, 5 (Items 14–17).
- Discussion: key results, limitations, interpretation, generalisability in main-text Sections 4.1–4.7 (Items 18–21).

#### S5. Detailed AI tool use disclosure

In accordance with BJPsych Open and Cambridge University Press research publishing ethics guidelines, the following is a full description of artificial intelligence (AI) tool use during the research and writing of this manuscript.

**S5.1 Tools used**

Claude (Anthropic), accessed through the claude.ai web interface. The model family used was Claude Sonnet/Opus.

**S5.2 Period of use**

AI-assisted code-drafting support was used during 2025–2026 while preparing analysis scripts.

**S5.3 Scope of use**

Claude was used only for code-drafting support, including drafting, refactoring and troubleshooting R/Python scripts used to run LDSC, LAVA, coloc and MAGMA-related workflows or to organise their outputs.

The AI tool was **not** used to:

- choose the study hypothesis;
- select the GWAS datasets;
- decide the statistical analysis plan;
- generate, alter or impute numerical results;
- interpret the findings;
- determine which results were significant;
- conduct independent literature review;
- write substantive scientific claims or conclusions.

**S5.4 Verification and accountability**

All scripts were reviewed, edited and executed by the author. Numerical outputs reported in the manuscript, figures and supplementary tables were cross-checked against the original analysis outputs. The author takes full responsibility for the integrity of the analyses, the accuracy of the reported values, the interpretation of results and the final manuscript.

**Figure S1. Regional association plot for ME/CFS-inf and IBS at chr12q24.22**

**Neither trait reached genome-wide significance ($p < 5\times10^{-8}$) at this locus; strongest signals for the two traits cluster in different sub-regions (coloc PP.H4 = 0.007)**

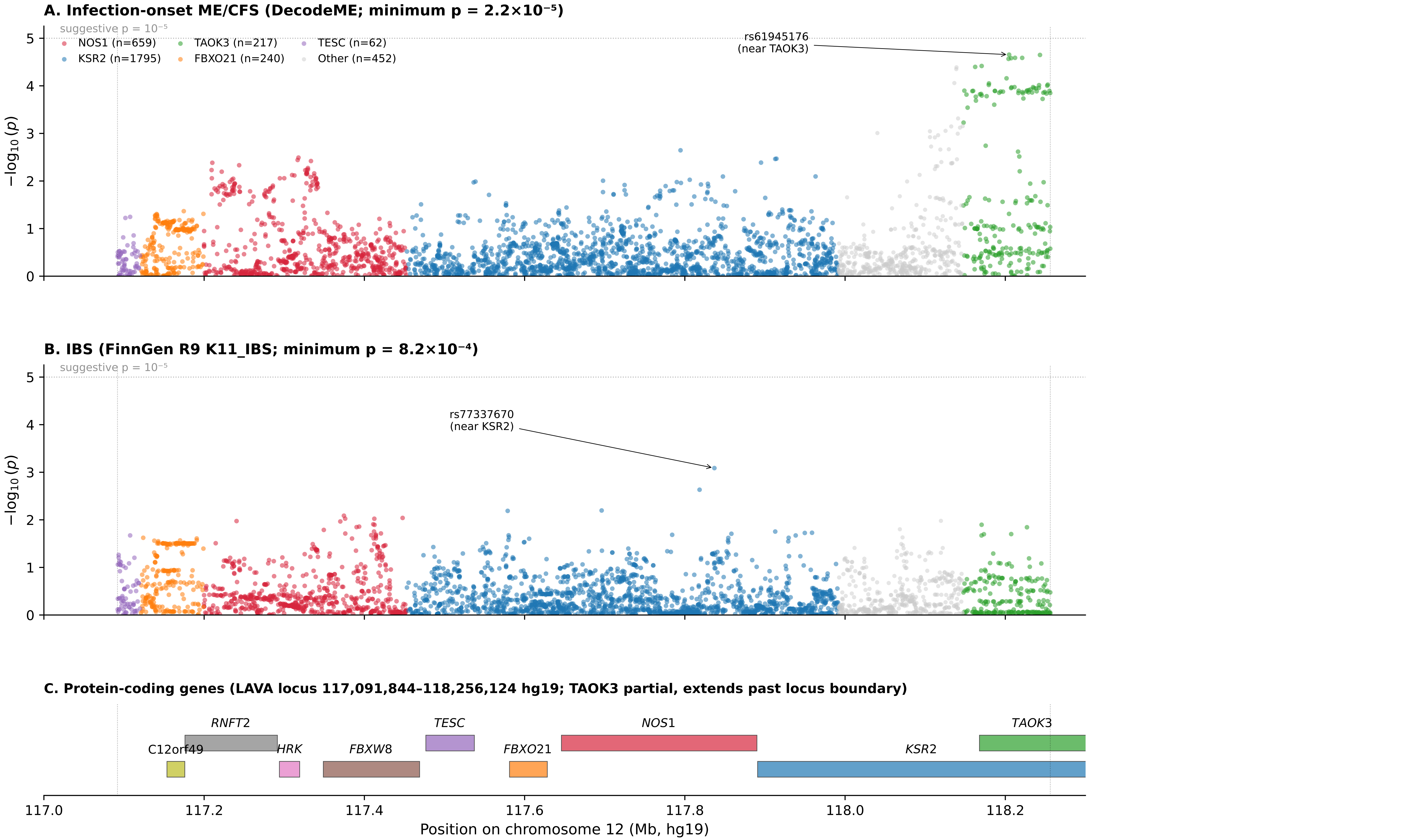
