## Supplementary material for "Global and local genetic overlap among ME/CFS, irritable bowel syndrome and psychiatric traits: a hypothesis-generating analysis": STREGA Checklist

**STREGA Reporting Checklist**

*Cross-trait local genetic correlation of infection-onset ME/CFS, irritable bowel syndrome, and psychiatric traits: a hypothesis-generating analysis*

Jun Hyun Lee

**Reporting framework.** This study reports a secondary analysis of publicly available GWAS summary statistics from the DecodeME, FinnGen R9, Psychiatric Genomics Consortium, and UK Biobank consortia, with the aim of estimating cross-trait global and local genetic correlations, single-cell heritability enrichment, and colocalisation. The STREGA (STrengthening the REporting of Genetic Association studies) extension of STROBE is the most applicable reporting framework. Because no new individual-level genotyping was performed, items relating to laboratory methods (e.g. genotyping platforms, allele calling, Hardy–Weinberg equilibrium testing, batch effects, relatedness) are addressed by the source consortia and cited accordingly; we have ensured that these items are reported transparently in the cited primary publications.

**Source publication references for genotyping methodology:** DecodeME (Devereux-Cooke et al., medRxiv 2025; doi:10.1101/2025.08.06.25333109), FinnGen R9 (Kurki et al., Nature 2023; 613:508–518), PGC MDD Wave 2 (Wray et al., Nat Genet 2018; 50:668–681), UK Biobank/IEU OpenGWAS (Elsworth et al., bioRxiv 2020; doi:10.1101/2020.08.10.244293).

| **Item No.** | **Domain** | **Recommendation** | **Reported in / location** |
| --- | --- | --- | --- |
| 1(a) | Title and abstract | Indicate the study's design with a commonly used term in the title or abstract. | Title ("cross-trait local genetic correlation… hypothesis-generating analysis"). Abstract Method paragraph names LDSC, LAVA, MAGMA, coloc. |
| 1(b) | Title and abstract | Provide an informative and balanced summary in the abstract of what was done and what was found. | Structured abstract: Background, Aims, Method, Results, Conclusions (248 words). |
| 2 | Introduction — background/rationale | Explain the scientific background and rationale for the investigation reported. | Section 1 (Introduction), paragraph 1: epidemiological link between post-infectious ME/CFS and IBS; paragraph 2: methodological rationale for local genetic correlation analysis. |
| 3 | Introduction — objectives | State specific objectives, including any pre-specified hypotheses. | Section 1, final paragraph: four explicit aims (i–iv). |
| 4 | Methods — study design | Present key elements of study design early in the paper. | Section 2.1 (Ethics) and 2.2 (Data sources): secondary cross-trait genetic correlation analysis of publicly available summary statistics. |
| 5 | Methods — setting | Describe the setting, locations, and relevant dates, including periods of recruitment, exposure, follow-up, and data collection. | Original consortium settings cited in Section 2.2 (DecodeME, FinnGen R9, PGC, UKB). Analysis period 2025–2026 (Section S5.2 of supplementary). |
| 6(a) | Methods — participants | Give the eligibility criteria and the sources and methods of case ascertainment and control selection. | Section 2.2 and Table 1: case/control definitions per consortium (ME/CFS: questionnaire-based diagnosis; IBS: ICD-10 K58 hospital registry codes; MDD: PGC MDD2 definitions; loneliness: UKB IEU continuous trait). |
| 6(b) | Methods — participants | For matched studies, give matching criteria and the number of controls per case. | Not applicable. Cases and controls matched at the original consortium level (see source publications). |
| 7(a) | Methods — variables | Clearly define all outcomes, exposures, predictors, potential confounders, and effect modifiers. Give diagnostic criteria, if applicable. | Section 2.2 and Table 1: outcome = case/control or continuous phenotype per trait. |
| 7(b) | Methods — variables (STREGA) | Clearly define genetic exposures (genetic variants) using a widely-used nomenclature system. | Section 2.5 (Colocalisation analysis): rsIDs (dbSNP), hg19 (GRCh37) coordinates. S-Table 8 reports rsID, chr, position, alleles. |
| 7(c) | Methods — variables (STREGA) | Identify variables likely to be associated with population stratification. | Section 2.2: all source datasets are European-ancestry. Population stratification controlled within each source GWAS by the original consortia. |
| 8(a) | Methods — data sources / measurement | For each variable of interest, give sources of data and details of methods of assessment. | Section 2.2; Table 1; references 2 (DecodeME), 8 (FinnGen), 9 (PGC MDD), 10 (UKB IEU). |
| 8(b) | Methods — measurement (STREGA) | Describe laboratory methods including source and storage of DNA, genotyping methods and platforms, error and call rates. | Reported in source consortium publications (DecodeME §2; FinnGen R9 Methods; PGC MDD2 Methods; UKB Methods). Not directly re-described, as no new genotyping was performed. |
| 8(c) | Methods — measurement (STREGA) | Specify whether genotypes were assigned using all of the data from the study simultaneously or in smaller batches. | Addressed by source consortia. All summary statistics analysed here are derived from imputed genotypes processed in single pipelines per cohort (HRC or 1000G reference panels; see cited sources). |
| 9 | Methods — bias | Describe any efforts to address potential sources of bias. | Section 2.3 (sample overlap via cross-trait LDSC intercept matrix), 2.7 (power considerations), 4.6 (Limitations). Supplementary S2.1 (QC) and S2.2 (LDSC intercept handling). |
| 10 | Methods — study size | Explain how the study size was arrived at. | Section 2.2 and Table 1: sample sizes determined by the originating consortia. Section 2.7: power considerations for the cross-trait analysis at the given sample sizes. |
| 11 | Methods — quantitative variables | Explain how quantitative variables were handled in the analyses. | Section 2.2: binary case/control coding for ME/CFS, IBS, MDD; continuous coding for loneliness/isolation (UKB ukb-b-8476). |
| 12(a) | Methods — statistical methods | Describe all statistical methods, including those used to control for confounding. | Sections 2.3 (LDSC), 2.4 (LAVA), 2.5 (Colocalisation), 2.6 (MAGMA). Supplementary S2.1–S2.5. |
| 12(b) | Methods — statistical methods | Describe any methods used to examine subgroups and interactions. | Section 2.2: pre-specified infection-onset subgroup (ME/CFS-inf) analysed separately from all-cases ME/CFS, with results compared in Sections 3.1–3.2. |
| 12(c) | Methods — statistical methods | Explain how missing data were addressed. | Section S2.1: SNPs with missing summary statistics excluded during munging; trait pairs filtered by LAVA univariate p<0.05 gating per locus. |
| 12(d) | Methods — sensitivity analyses | Describe any sensitivity analyses. | Section 2.5 (coloc sensitivity at p12 ∈ {1e-4, 1e-5, 1e-6}, S-Table 9). Section 2.6 (MAGMA top-10% binary mode as robustness check, S-Table 4). |
| 12(e) | Methods — software (STREGA) | State software version used and options (or settings) chosen. | Sections 2.3–2.6 and 2.8: LDSC v1.0.1, LAVA v0.1.5, coloc v5.2, MAGMA v1.10, GenomicSEM, R 4.3.x, Python 3.10.x. Supplementary S2 specifies non-default options (e.g. MAGMA TYPE=COVAR, direction=greater, condition-hide=Average). |
| 12(f) | Methods — Hardy-Weinberg (STREGA) | State whether Hardy-Weinberg equilibrium was considered and how. | Addressed by source consortia in their QC pipelines (cited consortium references). The HapMap3 SNP filter applied in munge_sumstats provides additional implicit HWE filtering. |
| 12(g) | Methods — imputation (STREGA) | Describe any methods used for inferring genotypes or haplotypes from observations or imputations. | Imputation performed by source consortia (DecodeME: HRC + UK10K; FinnGen: SISu v3; PGC MDD2: HRC; UKB: HRC + 1000G + UK10K). See source publications. |
| 12(h) | Methods — population stratification (STREGA) | Describe any methods used to assess or address population stratification. | Within-GWAS PCA-adjustment performed by source consortia. This study restricts to European-ancestry GWAS; cross-cohort population structure not directly comparable but controlled via LDSC's incorporation of LD scores from European 1000G reference (Section 2.3). |
| 12(i) | Methods — multiple testing (STREGA) | Describe any methods used to address multiple comparisons or to control risk of false positive findings. | Section 2.4: Benjamini–Hochberg FDR and Bonferroni applied across all 4,232 LAVA bivariate tests. MAGMA: within-trait FDR. LDSC: Bonferroni p<0.05 across 15 pairs. Discussed throughout Results. |
| 12(j) | Methods — relatedness (STREGA) | Describe any methods used to address and correct for relatedness among subjects. | Relatedness controlled within each source GWAS by the original consortia. Sample overlap across cohorts assessed via LDSC cross-trait intercept matrix (Section 2.3; S-Table 6). |
| 13(a) | Results — participants | Report numbers of individuals at each stage of study. | Table 1: per-trait sample sizes (cases, controls, total N). |
| 13(b) | Results — non-participation | Give reasons for non-participation at each stage. | Not applicable in a secondary analysis. Original consortia report participation/QC drop-out. |
| 13(c) | Results — flow diagram | Consider use of a flow diagram. | Workflow described narratively in Sections 2.3–2.6; a flow diagram is not provided but the analytic structure (5 traits × 9 trait pairs × 4,232 bivariate LAVA tests; 5 traits × 77 cell types in MAGMA; chr12q24.22 coloc with 3,425 SNPs) is fully specified. |
| 13(d) | Results — genotyping success (STREGA) | Report numbers of individuals in whom genotyping was attempted and were successful. | Addressed by source consortia. Numbers of SNPs surviving QC at each step of this analysis reported in Section 2.5 (coloc: 3,425 SNPs) and Section S2.1 (LDSC/LAVA: HapMap3 SNP set). |
| 14(a) | Results — descriptive data | Give characteristics of study participants. | Table 1: trait, case/control count, total N, observed-scale h². |
| 14(b) | Results — missing data | Indicate number of participants with missing data for each variable of interest. | Not applicable in a summary-statistics analysis at the participant level. SNP-level missingness handled at QC. |
| 14(c) | Results — by genotype (STREGA) | Consider giving information by genotype. | Not applicable; this analysis aggregates across genotypes within the summary-statistics framework. Effect sizes (β, SE) per SNP for the chr12 locus are provided in S-Table 8. |
| 15 | Results — outcome data | Report outcomes per genotype category over time. | Not applicable; cross-sectional summary-statistics design without longitudinal outcomes. |
| 16(a) | Results — main results with precision | Give unadjusted estimates and their precision (e.g., 95% CI). | Tables 2 (LDSC global rg with stated p), 3 (LAVA local correlations with FDR and Bonferroni), S-Tables 3 (95% CIs for ρ) and 5 (95% CIs for global rg). |
| 16(b) | Results — categorisation | Report category boundaries when continuous variables were categorized. | FDR and Bonferroni significance thresholds explicitly defined in Section 2.4 (FDR<0.05; Bonferroni p<1.2×10⁻⁵). |
| 16(c) | Results — multiple comparisons (STREGA) | Report results of any adjustments for multiple comparisons. | FDR (BH) and Bonferroni columns reported in Table 3 and S-Table 3. Within-trait FDR for MAGMA reported in S-Tables 4 and 7. |
| 17 | Results — other analyses | Report other analyses done — sensitivity analyses, replication. | Section 3.2: ME/CFS-all × IBS at chr12 (FDR=0.118, not significant — used as comparator). Section 3.3: top-10% MAGMA mode as robustness. Section S2.4: coloc sensitivity at 3 priors (S-Table 9). No independent GWAS replication of the chr12 locus was performed; this is acknowledged in Section 4.6. |
| 17(b) | Results — many tests summary (STREGA) | If numerous genetic exposures were examined, summarise results from all analyses undertaken. | All 4,232 LAVA bivariate tests reported in S-Table 3 (analytic set). All 11,520 univariate local heritability estimates in S-Table 1. All 385 MAGMA cell-type tests in S-Tables 4 and 7. |
| 18 | Discussion — key results | Summarise key results with reference to study objectives. | Section 4 (opening paragraph) and 4.7 (Summary). |
| 19 | Discussion — limitations | Discuss limitations, taking into account sources of bias or imprecision. | Section 4.6: seven explicit limitations (atlas resolution; borderline FDR finding; ICD-K58 phenotype; secondary analysis; observed-scale h²; self-reported infection-onset; no independent replication). |
| 20 | Discussion — interpretation | Give a cautious overall interpretation considering objectives, limitations, multiplicity, results from similar studies. | Sections 4.1–4.5 (NOS1 hypothesis, infection-onset specificity, MDD distinction, clinical implications, cell-type enrichment); 4.6 (limitations); 4.7 (summary). Throughout: framing as "hypothesis-generating". |
| 21 | Discussion — generalisability | Discuss the generalisability of the study results. | Section 4.6: limited to European-ancestry cohorts and FinnGen registry-defined IBS; cross-population transferability requires replication. Section 4.4: infection-onset stratification implications. |
| 22 | Other — funding | Give the source of funding and the role of funders. | "Funding" statement at end of main text: no specific funding received. Source-data acknowledgements in Acknowledgements. |

**Citation.** Little J, Higgins JPT, Ioannidis JPA, et al. STrengthening the REporting of Genetic Association studies (STREGA): an extension of the STROBE statement. *PLoS Med* 2009;6(2):e1000022.

**Notes.** Items relating to laboratory genotyping methods, Hardy–Weinberg equilibrium, imputation methodology, and within-cohort population stratification are addressed in the cited primary consortium publications (DecodeME, FinnGen R9, PGC MDD Wave 2, UK Biobank/MRC IEU). Items relating to longitudinal outcomes are not applicable to this cross-sectional summary-statistics design. All items necessary for transparent reproduction of the cross-trait analysis itself (statistical methods, software versions and settings, multiple-testing handling, sensitivity analyses, sample overlap correction, and reporting of all bivariate tests) are addressed in the main text and supplementary materials.
